# Genetic-epigenetic interactions (meQTLs) in orofacial clefts etiology

**DOI:** 10.1101/2025.02.09.25321494

**Authors:** AL Petrin, LA Machado-Paula, J Romanowska, RT Lie, L Hovey, B Doolittle, W Awotoye, L Dunlay, XJ Xie, E Zeng, A Butali, ML Marazita, JC Murray, LM Moreno-Uribe

## Abstract

**Objectives:** Nonsyndromic orofacial clefts (OFCs) involve complex genetic and environmental factors, with over 60 risk loci accounting for only a minority of estimated heritability and residing in non-coding regions with unclear functional relevance. We hypothesize that some genetic variants alter orofacial cleft risk by modifying DNA methylation (DNAm) at regulatory sequences essential for craniofacial development, acting as methylation quantitative trait loci (meQTLs).

**Methods:** We analyzed 10 well-established OFC-associated SNPs against genome-wide DNAm profiles in 409 cases and 456 controls, identifying 23 potential meQTLs. We validated findings using 358 cleft-discordant sibling pairs analyzed with quantitative MethyLight assays. Cross-referencing with the mQTL Database assessed temporal patterns across human development. Functional annotation used GeneHancer and craniofacial enhancer databases.

**Results:** Nine meQTLs were successfully replicated, including the highly significant rs987525 (8q24) - cg16561172 (*MYC*) association (P = 9.610E-6). This association mapped to a mesendoderm-active enhancer upstream of *MYC*, providing mechanistic explanation for the longstanding 8q24 cleft locus. Additional validated associations involved *MAFB-PLCG1, NOG-PPM1E, FOXE1-FRZB*, and *SPRY2-LGR4* interactions. Independent differential methylation analysis revealed significant differences between discordant siblings at three CpG sites. Cross-referencing confirmed concordance with population-level methylation effects, with childhood representing the critical developmental window for most associations.

**Conclusions:** This systematic meQTL characterization in OFCs demonstrates that genetic variants influence disease risk through epigenetic mechanisms. The 8q24-*MYC* regulatory pathway evidence provides crucial mechanistic insight into a major OFC risk locus. These findings bridge genetic associations with functional consequences, address missing heritability challenges, and suggest potential biomarkers and therapeutic targets for OFC prevention and treatment.

## Introduction

Nonsyndromic orofacial clefts (OFCs) affect approximately 1 in 700 live births worldwide and represent the most common craniofacial birth defects in humans(Rahimov, Jugessur et al. 2012). These complex malformations, classified as cleft lip with or without cleft palate (CL/P) and isolated cleft palate (CPO), arise from failures in normal embryonic fusion processes during the 6th-10th weeks of gestation. The clinical impact extends beyond the immediate structural defect, with affected individuals facing challenges in feeding, speech development, hearing, dental health, and psychosocial well-being throughout their lives(Fraser 1955; Marazita 2012).

The genetic architecture of OFCs exemplifies the complexity of common birth defects. While family history is observed in approximately 23% of cases indicating substantial genetic contribution, monozygotic twins show only 50% concordance rates(Grosen, Bille et al. 2010). This incomplete penetrance, combined with the observation that concordant twins often exhibit different phenotypic severity, suggests that factors beyond primary genetic risk variants contribute to disease manifestation and phenotypic variability.

Together, GWAS (Birnbaum, Ludwig et al. 2009; Grant, Wang et al. 2009; Beaty, Murray et al. 2010; Mangold, Ludwig et al. 2010; Camargo, Rivera et al. 2012; Sun, Huang et al. 2015; Wolf, Brand et al. 2015; Leslie, Liu et al. 2016), GWAS meta-analyses (Ludwig, Mangold et al. 2012; Beaty, Taub et al. 2013), linkage (Moreno, Mansilla et al. 2009) and replications studies have identified over 60 risk loci for OFCs; however, they account for a minority of the estimated risk and many of them reside in non-coding regions with unclear functional relevance so far. This challenge in translating statistical associations into biological mechanisms continues to limit our understanding of OFC etiology, necessitating expansion beyond single-omics analyses.

Recent advances in epigenomics have revealed DNA methylation as a critical mechanism through which genetic variants can influence gene expression and disease susceptibility. DNA methylation, involving the covalent addition of methyl groups to cytosine residues in CpG dinucleotides, serves as a key epigenetic modification regulating gene expression throughout development and adult life. Importantly, DNA methylation patterns can be influenced by both genetic variation and environmental factors, making them attractive candidates for explaining gene-environment interactions in complex diseases(Bourc’his, Xu et al. 2001; Song, Rechkoblit et al. 2011; van Eijk, de Jong et al. 2012).

Methylation quantitative trait loci (meQTLs) represent genetic variants that significantly influence DNA methylation levels at specific CpG sites. This framework provides a mechanistic bridge between genetic variation identified through GWAS and functional consequences at the molecular level. Large-scale studies in various tissues have demonstrated that genetic effects on DNA methylation are widespread, with recent analysis of over 27,000 blood samples showing that up to 45% of CpG sites are influenced by genetic variants(Villicaña and Bell 2021; Villicaña, Castillo-Fernandez et al. 2023).

The meQTL concept is particularly relevant for understanding non-coding GWAS variants, which comprise the majority of disease-associated loci. These variants may not directly alter protein-coding sequences but instead influence regulatory elements that control gene expression through epigenetic mechanisms(Villicaña, Castillo-Fernandez et al. 2023). For OFCs, where many risk variants are located in non-coding regions near developmental genes, meQTL analysis offers a systematic approach to identify functional mechanisms.

Emerging evidence supports that differential DNAm may alter risk for different cleft types and modify OFCs penetrance (Alvizi, Ke et al. 2017). Our recent work showed that differential methylation contributes to phenotypic variability in patients with Van der Woude syndrome, including monozygotic twins with discordant phenotypes despite carrying identical causal mutations in the *IRF6* gene (Petrin, Zeng et al. 2023b; Seaberg, Awotoye et al. 2024). Some studies indicate that epigenetic factors such as DNA methylation (DNAm) are associated with OFCs risk (Joubert, Felix et al. 2016; Alvizi, Ke et al. 2017; Sharp, Ho et al. 2017; Shu, Shu et al. 2018; Gonseth, Shaw et al. 2019; Xu, Lie et al. 2019; Young, Slifer et al. 2021; Alvizi, Brito et al. 2022; Charoenvicha, Sirimaharaj et al. 2022; Zhang, Zhang et al. 2023), Moreover, differentially methylated regions tend to cluster around gene pathways previously linked to palatogenesis, and that some of these differential DNAm regions seem to be influenced by genetic variation (Alvizi, Ke et al. 2017; Sharp, Ho et al. 2017; Howe, Richardson et al. 2018; Howe, Richardson et al. 2019). However, systematic characterization of meQTLs specifically in the context of OFC-associated genetic variants has not been performed, representing a significant knowledge gap in understanding the functional mechanisms underlying genetic risk for these common birth defects.

Therefore, analyses that combine known genetic risk factors with epigenetic data to explore meQTLs function significantly contribute to understanding such loci by addressing the causal alleles, their pathogenic consequences, and the biological mechanisms through which they influence OFCs risk.

Discordant sibling pairs are valuable cohorts for mapping complex human traits. The use of discordant sibling pairs in methylation studies offers several advantages: siblings share a significant portion of their genetic makeup, which helps control for genetic variation and isolates environmental influences on methylation. Additionally, siblings often experience similar early-life environments, aiding in the study of environmental impacts on DNA methylation(Kim, Kwak et al. 2017). Comparing affected and unaffected siblings (discordant pairs) can reveal disease-related methylation changes, and the reduced variability from genetic and environmental similarities enhances the statistical power and reliability of the study.

## Methods

### Study Design and Samples

Our study employed a two-stage design to identify and validate meQTLs associated with orofacial clefts. The discovery cohort (Cohort 1) consisted of unrelated individuals with genome-wide methylation data, while the validation cohort (Cohort 2) comprised cleft-discordant sibling pairs to control for genetic background and shared environmental factors.

#### Cohort 1 (Discovery)

We analyzed a subset of unrelated case with cleft lip with or without cleft palate and unaffected controls from previous published studies (Moreno Uribe, Fomina et al. 2017; Romanowska, Haaland et al. 2020). We obtained genotype data from 10 cleft-associated SNPs selected based on their identification as top loci for OFCs in previous GWAS and candidate gene studies, along with genome-wide DNA methylation data (obtained with Illumina 450K array) for 409 cases with OFCs and 456 controls, all with DNA extracted from blood.

#### Cohort 2 (Validation)

The replication cohort consisted of 358 pairs of same-sex siblings discordant for cleft lip with or without cleft palate with DNA from blood (n=164 pairs) and DNA from saliva (n=194 pairs) (Table 1). All samples were obtained as part of previous studies following approval by the University of Iowa and local Institutional Review Boards (IRBs). Informed consent was provided by patients, parents, or guardians prior to sample collection and clinical information acquisition.

**Table 1:** Distribution of samples.

| Characteristic | Total Samples (N) / Sib pairs (N) | % |
| --- | --- | --- |
| **Total Samples** | 716/358 | 100% |
| **Gender** |  |  |
| Male | 364/182 | 51% |
| Female | 352/176 | 49% |
| **Sample Source** |  |  |
| Blood | 328/164 | 46% |
| Saliva | 388/194 | 54% |
| **Cleft Type** |  |  |
| CL | 80 | 22% |
| CLP | 254 | 71% |
| CPO | 24 | 7% |
| **Population** |  |  |
| Asia | 238 | 33.2% |
| South America | 188 | 26.2% |
| North America | 146 | 20.0% |
| Africa | 60 | 8.0% |
| Europe | 48 | 7.0% |
| Central America | 36 | 5.0% |

N= number of samples; CL=Cleft lip; CLP= Cleft lip and palate; CPO= Cleft palate only

### Genotyping

We selected 10 SNPs representing well-established OFC risk loci based on their identification as genome-wide significant associations in previous GWAS and meta-analyses(Romanowska, Haaland et al. 2020) (Moreno Uribe, Fomina et al. 2017) (Table 2). These SNPs were chosen to represent the strongest and most replicated genetic associations with OFCs, spanning different chromosomes and putative functional mechanisms.

**Table 2:** Most significant meQTLs resulting from case-control study.

| SNP | SNP gene | CpG | CpG target gene | P value |
| --- | --- | --- | --- | --- |
| rs4752028 | VAX1 | cg08319991 | UCHL1 | 2.83E-08 |
| rs8001641 | SPRY2 | cg19191560 | LGR4 | 2.88E-08 |
| rs7590268 | THADA | cg06873343 | N/A | 7.72E-08 |
| rs7078160 | VAX1 | cg08319991 | UCHL1 | 1.11E-07 |
| rs3758249 | FOXE1 | cg20308679 | FRZB | 1.29E-07 |
| rs4752028 | VAX1 | cg11876012 | AFAP1L2 | 1.41E-07 |
| rs227731 | NOG | cg08592707 | SKA2 | 1.59E-05 |
| rs1873147 | TPM1 | cg11936410 | TPM1 | 5.41E-04 |
| rs560426 | ABCA4/ARHGAP29 | cg25196715 | BCAR3/MIR760 | 6.72E-04 |
| rs987525 | 8q24 | cg16561172 | MYC | 7.04E-04 |
| rs560426 | *ABCA4/ARHGAP29* | cg00405232 | *ABCA4* | 7.93E-04 |
| *rs227731* | *NOG* | cg01964121 | *PPM1E* | 8.11E-04 |
| *rs227731* | *NOG* | cg10303698 | *CUEDC1* | 8.74E-04 |
| rs1873147 | *TPM1* | cg24483493 | *HERC1* | 9.14E-04 |
| rs4752028 | *VAX1* | cg08329473 | *AFAP1L2* | 9.29E-04 |
| rs13041247 | *MAFB* | cg18347630 | *CHD6* | 1.02E-03 |
| *rs7078160* | *VAX1* | cg08329473 | *AFAP1L2* | 1.06E-03 |
| rs1873147 | *TPM1* | cg16659880 | *TPM1* | 1.11E-03 |
| rs7078160 | *VAX1* | cg09487139 | *N/A* | 1.38E-03 |
| rs13041247 | *MAFB* | cg00514723 | *LPIN3* | 1.39E-03 |
| rs742071 | *PAX7* | cg20940024 | *N/A* | 1.56E-03 |
| rs1873147 | *TPM1* | cg19631779 | *TPM1* | 1.67E-03 |
| rs13041247 | *MAFB* | cg17103269 | *PLCG1* | 1.78E-03 |

SNPs in italic indicate SNPs that have an meQTL role in other tissues or phenotypes

For cohort 1, we extracted the genetic data for the selected 10 SNPs (Table 2) from previous published studies (Romanowska, Haaland et al. 2020) (Moreno Uribe, Fomina et al. 2017). Detailed information regarding data collection, quality control, and preprocessing is available in the appendix of the cited work. For cohort 2, we genotyped the 10 SNPs in discordant sibling pairs using Taqman assays on a Fluidigm nanofluidic platform (Fluidigm Corp., South San Francisco, CA, USA). Genotype calling was performed using Fluidigm SNP genotyping software version 4.1.2 with default settings. Quality control included setting the confidence threshold to 65% for the genotype calling algorithm followed by visual inspection of all genotyping plots.

### Bisulfite conversion and DNA methylation profiling

All samples were bisulfite converted prior measuring DNA methylation levels. Briefly, DNA quality was assessed QubitTM dsDNA High Sensitivity Range Assay Kit (Thermo Fisher Scientific) and 1.5% agarose gel. After quantification of each sample, 500 ng of each genomic DNA sample was submitted to bisulfite conversion using the EZ DNA Methylation Kit (Zymo Research) according to manufacturer’s protocol.

For **cohort 1**, DNA methylation levels were measured using the Illumina Human Methylation 450K BeadChip (Illumina, Inc., San Diego, CA). DNA methylation level was then estimated at 485,577 CpG sites and the HAPLIN R package was used to extract the raw probe intensity values. These raw values were preprocessed using ENmix R package(Xu, Niu et al. 2021). The following criteria were used to identify low-quality samples: (1) average intensity value across internal control probes less than 5500, (2) more than 5% of CpG probes having low-quality data (Illumina detection P value > 10^−6^, read from less than 3 beads, or outlier value for the probe in the dataset), and (3) clear outliers based on visual inspection of a density plot of total intensity. Next, the low quality CpG probes were defined as follows: (1) more than 5% low-quality data; (2) common SNP within the probe’s sequence (minor allele frequency > 0:05 in Europeans based on 1000 Genomes Project data), or probes mapping to multiple genomic locations, or CpGs on X or Y chromosomes; (3) CpGs with multiple mode distributions identified with ENmix. More details about the filtering steps can be found in the cited work(Romanowska, Haaland et al. 2020).

For **cohort 2**, after bisulfite conversion, genomic DNA samples were amplified by fluorescence-based, real time quantitative PCR (Methylight)(Eads, Danenberg et al. 2000). Quantitative MethyLight assays (e.g., EpiTect MethyLight Assays) consist of two probes, one methylation specific and the other nonmethylation specific, which can be used in a single real-time PCR reaction. This enables highly accurate quantitative methylation analysis, due to the simultaneous detection of methylated and unmethylated DNA. We used locus-specific PCR primers flanking an oligonucleotide probe with a 5’ fluorescent reporter dye to detect methylated (SUN) and unmethylated (FAM) alleles and a 3’ quencher dye (3IABkFQ). The primers and fluorescent probes were designed against bisulfite-converted DNA sequence and quantitative information was obtained in real time. Serial dilutions of the EpiTect Control DNAs (Qiagen) were included on each plate to generate a standard curve and to verify plate to plate consistency. The PCR amplification was performed in a 384-well plate format and each sample was ran in duplicate. EpiTect MethyLight assays enable the direct quantification of the methylation degree in a sample by taking the threshold cycles (C_T_) determined in the SUN channel with the probe detecting methylated DNA or in the FAM channel with the probe detecting unmethylated DNA. Ten nanograms of bisulfite converted, methylated and unmethylated human control DNA, or defined mixtures of both DNAs were used for methylation quantification. The methylation degree of each sample was calculated from the average of C_T_ values in SUN (C_T(CG)_) and FAM (C_T(TG)_) channel, obtained in quantitative real-time PCR using the formula described in (Cottrell, Jung et al. 2007): C_meth_ = 100/[1+2^(C^_T(CG)_ ^−C^_T(TG)_^)^]%; where C_T(CG)_ is the methylated signal obtained by the threshold cycle of the CG reporter (SUN channel), and C_T(TG)_ is the unmethylated signal obtained by the threshold cycle of the CG reporter (FAM channel).

### Statistical analysis

#### Discovery Analysis (Cohort 1)

We performed meQTL analysis using the R package MatrixEQTL(Shabalin 2012), testing associations between each of the 10 SNPs and all 407,513 high-quality CpG sites that passed QC. Linear regression models assessed the relationship between CpG methylation levels and SNP genotype (additive model: 0, 1, or 2 copies of the minor allele), using cleft status as a covariate.

The statistical model was *Methylation ~ Genotype + Cleft_Status + Covariates*. We applied Benjamini-Hochberg false discovery rate (FDR) correction for multiple testing, with significance defined as P < 0.002 and FDR < 0.4 (Table 3). The FDR threshold of 0.4 was chosen to balance discovery sensitivity with replication feasibility, consistent with approaches used in large-scale meQTL studies.

**Table 3:** Replicated SNP-CpG associations (meQTLs) from linear analysis in all groups.

| Sample | Affected/<br>Unaffected | SNPs | SNP gene | CpG | CpG gene | P value |
| --- | --- | --- | --- | --- | --- | --- |
| All | 353/353 | rs13041247 | *MAFB* | cg18347630 | *PLCG1* | 2.13E-01 |
| Blood | 160/160 | rs13041247 |  | cg18347630 |  | **3.93E-02** |
| Saliva | 193/193 | rs13041247 |  | cg18347630 |  | 3.38E-01 |
| All | 354/354 | rs227731 | *NOG* | cg08592707 | *PPM1E* | **1.54E-02** |
| Blood | 163/163 | rs227731 |  | cg08592707 |  | 3.66E-01 |
| Saliva | 191/191 | rs227731 |  | cg08592707 |  | 5.34E-01 |
| All | 300/300 | rs227731 | *NOG* | cg10303698 | *CUEDC1* | **1.50E-03** |
| Blood | 110/110 | rs227731 |  | cg10303698 |  | 5.08E-01 |
| Saliva | 190/190 | rs227731 |  | cg10303698 |  | **5.95E-02** |
| All | 318/318 | rs3758249 | *FOXE1* | cg20308679 | *FRZB* | 1.10E-01 |
| Blood | 131/131 | rs3758249 |  | cg20308679 |  | 2.92E-01 |
| Saliva | 187/187 | rs3758249 |  | cg20308679 |  | **3.80E-02** |
| All | 340/340 | rs8001641 | *SRPY2* | cg19191560 | *LGR4* | **4.70E-02** |
| Blood | 158/158 | rs8001641 |  | cg19191560 |  | **9.33E-02** |
| Saliva | 182/182 | rs8001641 |  | cg19191560 |  | **3.72E-02** |
| All | 305/305 | rs987525 | *8q24* | cg16561172 | *MYC* | **2.78E-05** |
| Blood | 138/138 | rs987525 |  | cg16561172 |  | 6.12E-01 |
| Saliva | 167/167 | rs987525 |  | cg16561172 |  | **9.63E-06** |
| All | 356/356 | rs7590268 | *THADA* | cg06873343 | *TTYH3* | 3.08E-01 |
| Blood | 165/165 | rs7590268 |  | cg06873343 |  | **3.91E-02** |
| Saliva | 191/191 | rs7590268 |  | cg06873343 |  | 9.14E-01 |
| All | 333/333 | rs7078160 | *VAX1* | cg09487139 | N/A | 7.30E-01 |
| Blood | 157/157 | rs7078160 |  | cg09487139 |  | **4.60E-02** |
| Saliva | 176/176 | rs7078160 |  | cg09487139 |  | 2.75E-01 |
| All | 297/297 | rs560426 | *ABCA4/*<br>*ARHGAP29* | cg25196715 | *ABCA4/*<br>*ARHGAP29* | **3.16E-02** |
| Blood | 126/126 | rs560426 |  | cg25196715 |  | 3.88E-01 |
| Saliva | 171/171 | rs560426 |  | cg25196715 |  | **3.81E-02** |

P values in bold are borderline significant (P<0.09); FDR=False Discovery Rate
Beta value ranges: 0-0.2 = fully unmethylated; 0.21-0.60 = partially methylated; 0.61-1.00 = fully methylated

#### Validation Analysis (Cohort 2)

We tested the 23 candidate SNP-CpG associations identified in Cohort 1 using the the 358 discordant sibling pairs and the same statistical framework in MatrixEQTL. To assess methylation differences independent of SNP effects, we performed paired Student’s t-tests comparing methylation levels between affected and unaffected siblings within each discordant pair for each tissue subgroup (blood, saliva, and combined).

#### Functional Annotation

We investigated the regulatory potential of validated meQTL-associated CpG sites using the GeneHancer database(Fishilevich, Nudel et al. 2017), — a regulatory element database containing genomic coordinates of known enhancer and promoter elements, including those active during craniofacial development (Wilderman, VanOudenhove et al. 2018; Yankee, Oh et al. 2023). We compared these coordinates to our CpG sites to: (1) investigate whether validated CpG sites overlapped with regulatory elements and (2) prioritize the most likely CpG-target genes in relevant tissues (e.g., epithelial and mesenchymal tissues) based on GeneHancer *in silico* prediction tools.

To further support the potential role of the selected SNPs as meQTLs, we searched the mQTL Database (http://www.mqtldb.org/) (Gaunt, Shihab et al. 2016) for a systematic identification of genetic influences on methylation across the serial time points on human blood samples. This database contains datasets of large-scale genome-wide DNA methylation analysis of 1,000 mother-child pairs; and presents a comprehensive genome-wide cis and trans meQTL longitudinal analysis in blood DNA at three time points in their life course.

## Results

Our two-stage study design included a discovery cohort (Cohort 1) of 865 unrelated individuals (409 OFC cases and 456 controls) and a validation cohort (Cohort 2) of 358 cleft-discordant sibling pairs, totaling 716 individuals (Table 1). The combined cohorts showed balanced sex distribution (51% male, 49% female) and representation across multiple populations, with Asian populations comprising the largest group (33.2%), followed by South American (26.2%), North American (20%), African (8%), European (7%), and Central American (5%) populations. DNA samples were obtained from blood in 46% of cases and saliva in 54% of cases. Among affected individuals, cleft lip and palate (CLP) represented the most common phenotype (71%), followed by isolated cleft lip (CL, 22%) and isolated cleft palate (CPO, 7%).

### Discovery of meQTL Associations in the Case-Control Cohort

Analysis of 10 well-established OFC-associated SNPs against 407,513 high-quality CpG sites in the discovery cohort identified 23 significant SNP-CpG associations meeting our criteria (P < 0.002, FDR < 0.4) (Table 2). The strongest association was observed between rs4752028 (*VAX1*) and cg08319991 (*UCHL1*) with P = 2.83 E-08, followed closely by rs8001641 (*SPRY2*) and cg19191560 (*LGR4*) with P = 2.88 E-08. The third most significant association involved rs7590268 (*THADA*) and cg06873343 (*TTYH3*) at P = 7.72 E-08.

Several associations involved the same SNPs affecting multiple CpG sites, suggesting pleiotropic regulatory effects. The rs4752028 (*VAX1*) variant was associated with both cg08319991 (*UCHL1*, P = 2.83 E-08) and cg11876012 (*AFAP1L2*, P = 1.41 E-07), indicating that this variant may influence multiple downstream targets. Similarly, rs227731 (*NOG*) showed associations with multiple CpG sites including cg08592707 (*SKA2*, P = 1.59 E-05) and cg01964121 (*PPM1E*, P = 8.11 E-04). The well-known 8q24 variant rs987525, representing one of the most replicated OFC risk loci across populations, showed significant association with cg16561172 (*MYC*) at P = 7.04 E-04.

The identified meQTL associations included both cis-acting effects, where SNPs and CpG sites were located on the same chromosome or in proximity, such as rs1873147 (*TPM1*) with cg11936410 (*TPM1*) and rs560426 (*ABCA4/ARHGAP29*) with cg00405232 (*ABCA4*), as well as trans-acting effects across chromosomes, indicating long-range regulatory mechanisms that may involve chromatin looping or other three-dimensional nuclear architecture.

### Validation of meQTL Associations in Cleft-Discordant Sibling Pairs

Of the 23 candidate meQTL associations identified in the discovery phase, 9 were successfully replicated in the independent cohort of cleft-discordant sibling pairs (Table 3). The validation analysis revealed distinct tissue-specific patterns and combined effects across blood and saliva samples.

In blood samples, three meQTL associations achieved statistical significance. The interaction between rs13041247 (*MAFB*) and cg18347630 (*PLCG1*) reached significance at P = 3.93 E^−^02, while rs7590268 (*THADA*) showed association with cg06873343 (*TTYH3*) at P = 3.91E-02. Additionally, rs7078160 (*VAX1*) demonstrated association with cg09487139 at P = 4.60E-02, though this CpG site did not map to a specific gene region.

Analysis of saliva samples revealed distinct tissue-specific patterns. The rs3758249 (*FOXE1*) variant showed significant association with cg20308679 (*FRZB*) at P = 3.80E-02, representing an important connection between two genes previously linked to OFC development. The rs8001641 (*SPRY2*) variant was associated with cg19191560 (*LGR4*) at 3.72E-02. Most remarkably, rs987525 (8q24) demonstrated a highly significant association with cg16561172 (*MYC*) at P = 9.63E-06, representing the strongest statistical signal in our entire validation analysis. The rs560426 (*ABCA4/ARHGAP29*) variant showed association with cg25196715 (*ABCA4/ARHGAP29*) at 3.81E-02, demonstrating a cis-acting regulatory effect.

When blood and saliva samples were analyzed together, several associations achieved statistical significance. The rs227731 (*NOG*) variant showed associations with two distinct CpG sites, cg08592707 (*PPM1E*) with P = 1.54E-02 and cg10303698 (*CUEDC1*) with P = 1.50E-03, with the latter representing one of the most significant findings in the validation cohort. The rs8001641 (*SPRY2*) association with cg19191560 (*LGR4*) remained significant at P = 4.70E-02 in the combined analysis. The 8q24 association remained highly robust, with rs987525 and cg16561172 (*MYC*) showing P = 2.78E-05 in the combined analysis. The rs560426 (*ABCA4/ARHGAP29*) association with cg25196715 (*ABCA4/ARHGAP29*) achieved P = 3.16E-02 in the combined analysis.

The most statistically robust finding across all analyses was the rs987525 (8q24) association with cg16561172 (*MYC*), which achieved genome-wide significance in saliva samples and remained highly significant in the combined analysis, providing compelling evidence for a functional mechanism underlying this well-established but poorly understood OFC risk locus.

### Differential DNA Methylation Analysis Between Discordant Siblings

Independent of SNP effects, paired t-test analyses comparing methylation levels between affected and unaffected siblings revealed significant differential methylation at specific CpG sites (Table 4). The cg06873343 (*TTYH3*) site showed the strongest and most consistent differential methylation signal across all analyses. In the combined analysis of 358 sibling pairs, this site achieved P = 4.17E-03, with consistent significance maintained in blood-specific analysis of 165 pairs (P = 3.76E-02) and saliva-specific analysis of 193 pairs (P = 4.94E-02). This remarkable consistency across different tissue types strongly indicates robust differential methylation at this locus that is independent of tissue-specific factors.

**Table 4:**
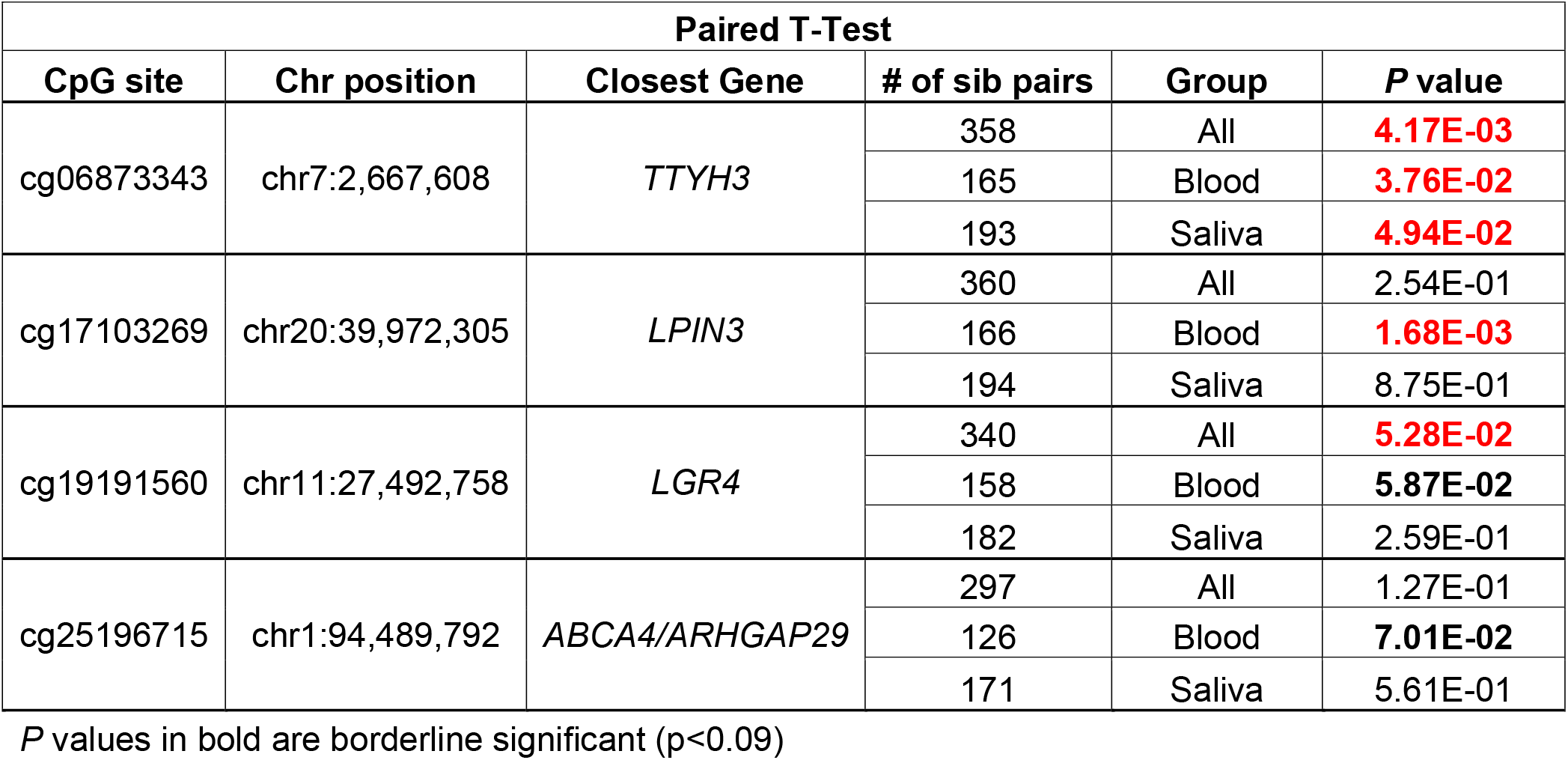
Differential methylation levels between sibling pairs and subdivided by groups.

The cg17103269 (*LPIN3*) site demonstrated clear blood-specific differential methylation with high statistical significance in blood samples from 166 pairs (P = 1.68E-03), representing one of the strongest signals in our differential methylation analysis. However, this effect was not observed in saliva samples from 194 pairs (P = 8.75E-01) or in the combined analysis of 360 pairs (P = 2.54E-01), suggesting tissue-specific regulatory mechanisms.

The cg19191560 (*LGR4*) site showed borderline significance in the combined analysis of 340 pairs (P = 5.28E-02), with similar trends observed in blood-specific analysis of 158 pairs (P = 0.059) but not in saliva-specific analysis of 182 pairs (P = 2.59E-01). The cg25196715 (*ABCA4/ARHGAP29*) site showed suggestive evidence for differential methylation, particularly in blood samples from 126 pairs (P = 7.01E-02), though this did not reach conventional statistical significance thresholds.

### Functional Annotation of Validated meQTL Sites

Analysis using the GeneHancer and craniofacial enhancers databases (Wilderman, VanOudenhove et al. 2018; Yankee, Oh et al. 2023) revealed that several validated CpG sites overlapped with known regulatory elements active during craniofacial development. The cg16561172 site associated with rs987525 (8q24) mapped to a mesendoderm-active enhancer region (ENSR8_CGN7Z) located upstream of *MYC*, providing the first mechanistic insight into how this long-standing mystery locus may influence OFC risk through epigenetic regulation of this critical developmental gene. The cg08592707 site linked to rs227731 (*NOG*) resided within a promoter region (ENSR17_9TBN3) active in dermal fibroblasts, mesenchymal stem cells, and mesendoderm, which is consistent with its association with *PPM1E*, a gene known to be involved in osteoblast proliferation and bone formation processes critical for proper craniofacial development.

The cg20308679 site associated with rs3758249 (*FOXE1*) mapped to a promoter region (ENSR2_94NKPT) active in dermal fibroblasts, foreskin fibroblasts, mesenchymal stem cells, and mesendoderm. This regulatory element annotation supports the biological relevance of the FOXE1-FRZB interaction, as both genes have established roles in craniofacial development and the Wnt signaling pathway.

Cross-referencing our findings with the mQTL Database confirmed that seven of our identified SNPs function as meQTLs in other tissues and developmental contexts, supporting the validity of our findings while highlighting the tissue-specific and context-dependent nature of some associations (Table 5). This functional annotation analysis supports the biological plausibility of our identified meQTL associations and provides evidence that they represent genuine regulatory interactions rather than spurious statistical associations, establishing a foundation for future functional validation studies.

**Table 5:**
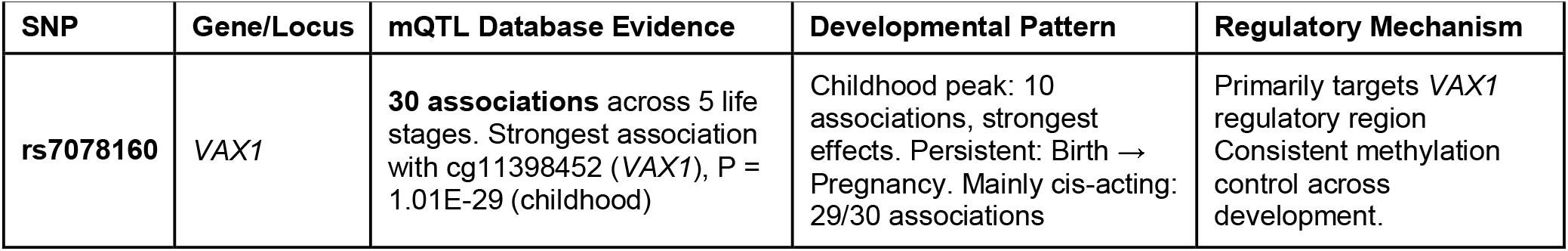
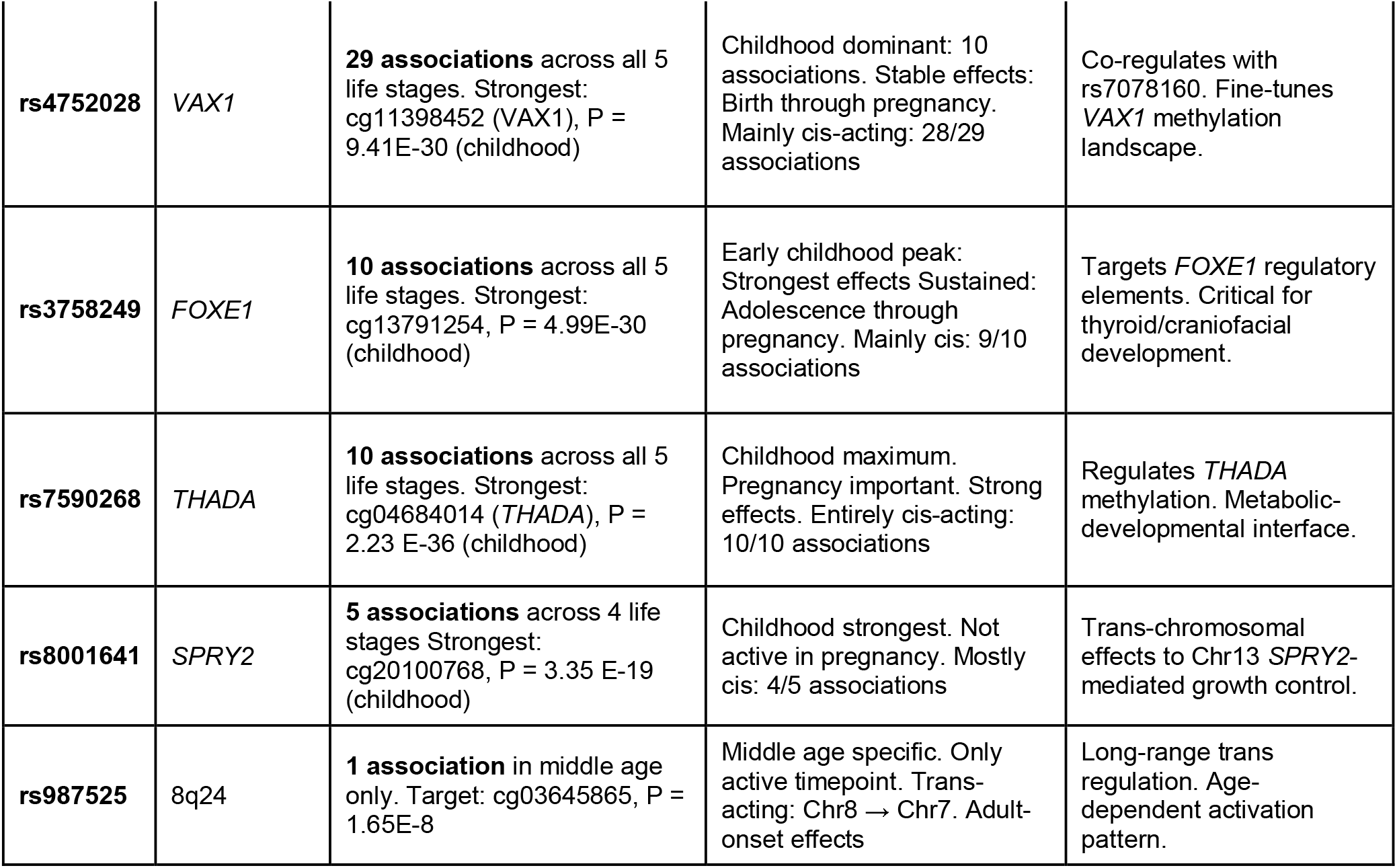
Functional Analysis of SNPs Functioning as meQTLs in Other Tissues and Developmental Contexts.

## Discussion

This study represents the first systematic investigation of methylation quantitative trait loci (meQTLs) specifically in the context of orofacial clefts, addressing a critical gap in understanding how genome-wide association study (GWAS) variants influence disease risk through epigenetic mechanisms (Villicaña and Bell 2021; Villicaña, Castillo-Fernandez et al. 2023). Our two-stage approach successfully identified and validated nine meQTL associations between established OFC risk variants and DNA methylation sites, providing mechanistic insights that bridge genetic variation with functional consequences during craniofacial development.

The most significant finding of our study concerns the long-standing mystery surrounding the 8q24 locus and its association with OFC risk. The rs987525 variant at 8q24 has been consistently identified across multiple GWAS as one of the strongest genetic risk factors for orofacial clefts (Birnbaum, Ludwig et al. 2009; Grant, Wang et al. 2009; Beaty, Murray et al. 2010; Ludwig, Mangold et al. 2012; Salagovic, Klimcakova et al. 2017; Butali, Mossey et al. 2019) yet the functional mechanism underlying this association has remained elusive for over a decade. Our meQTL analysis provides compelling mechanistic explanation for this association by evidencing that rs987525 significantly modulates methylation at cg16561172, located upstream of the *MYC* gene (P = 9.6 × 10E-6). This methylation site overlaps with a mesendoderm-active enhancer region (Fishilevich, Nudel et al. 2017; Wilderman, VanOudenhove et al. 2018; Yankee, Oh et al. 2023), suggesting that the 8q24 risk allele disrupts normal epigenetic regulation of *MYC* during critical developmental windows. The *MYC* gene encodes a master regulator of cellular proliferation, growth, and differentiation (Meyer and Penn 2008; Dang 2012), processes that are fundamentally critical during craniofacial morphogenesis (Trainor 2014; Twigg and Wilkie 2015). Even subtle dysregulation of MYC expression could lead to defects in palatal shelf growth, fusion timing, or both, explaining how this distant regulatory variant influences OFC susceptibility through long-range epigenetic control (Karsenty and Wagner 2002; Long and Ornitz 2013).

The unique temporal pattern of rs987525 in the mQTL Database (Gaunt, Shihab et al. 2016)showing methylation effects exclusively during middle age through trans-chromosomal regulation, suggests this variant may have pleiotropic effects across the lifespan. This age-specific activation pattern distinguishes the 8q24 locus from other OFC variants and may explain why this region has been difficult to characterize functionally using traditional developmental biology approaches that focus on embryonic stages (Santagati and Rijli 2003; Chai and Maxson 2006). The finding that this variant exerts its strongest effects through adult methylation patterns indicates that the 8q24 locus may influence OFC risk through maternal epigenetic factors or transgenerational inheritance mechanisms (Gapp, Jawaid et al. 2014; Heard and Martienssen 2014), opening new avenues for understanding gene-environment interactions in cleft etiology (Waterland and Michels 2007; Heijmans, Tobi et al. 2008).

Our cross-referencing with longitudinal methylation data (mQTL Database (Gaunt, Shihab et al. 2016)) revealed that OFC-associated variants primarily exert their methylation effects during childhood, aligning with the critical periods of craniofacial development (Sperber, Saxena et al. 2008; Marazita 2012). Five of the six validated SNPs (rs7078160, rs4752028, rs3758249, rs7590268, and rs8001641) showed their strongest methylation associations during childhood, with effect sizes reaching genome-wide significance levels. This temporal clustering provides compelling evidence that these genetic variants influence OFC risk by modulating epigenetic patterns during developmentally relevant windows rather than through constitutive effects across all life stages (Burdge and Lillycrop 2010; Waterland, Kellermayer et al. 2010).

The tissue-specific patterns observed in our validation cohort further highlight the complexity of epigenetic regulation in OFC etiology (Alvizi, Ke et al. 2017; Petrin, Zeng et al. 2023a). The differential effects observed between blood and saliva samples suggest that genetic variants may influence methylation patterns differently across tissue types (Pai, Bell et al. 2011; Lokk, Modhukur et al. 2014).

Our meQTL analysis revealed associations involving genes not traditionally linked to craniofacial development, expanding the known molecular landscape underlying OFC etiology. The identification of significant associations involving *TTYH3* (chloride channel) (Suzuki and Mizuno 2004; Halleran, Sehdev et al. 2015), *PPM1E* (osteoblast proliferation regulator) (Li, Zhu et al. 2017; Kanazawa, Takeno et al. 2018), *PLCG1* (cell growth and migration) (Tao, Han et al. 2023), and *LGR4* (Wnt signaling modulator) (de Lau, Barker et al. 2011; Glinka, Dolde et al. 2011) suggests that OFC susceptibility extends beyond canonical palatogenesis pathways to encompass broader cellular processes including ion transport, metabolic regulation, and immune homeostasis (Sauka-Spengler and Bronner-Fraser 2008; Theveneau and Mayor 2012).

The *TTYH3* association is particularly noteworthy given its consistent differential methylation across tissue types and its meQTL interaction with *THADA. TTYH3* encodes a large-conductance chloride channel with expression during embryonic nervous system development, suggesting potential roles in neural crest cell migration or epithelial-mesenchymal transitions during facial morphogenesis (Thiery, Acloque et al. 2009; Nieto, Huang et al. 2016). The *PPM1E* association links OFC risk to osteoblast proliferation control, providing a direct connection to bone formation processes critical for proper craniofacial structure development (Karsenty, Kronenberg et al. 2009; Long 2011).

These findings suggest potential therapeutic avenues that might not have been evident through genetic analysis alone. Targeting DNA methylation patterns through epigenetic therapies could theoretically modify OFC risk, particularly during critical developmental windows (Sharma, Kelly et al. 2010; Dawson and Kouzarides 2012). However, such approaches would require careful consideration of timing, tissue specificity, and potential off-target effects given the fundamental roles these pathways play in normal development (Sharma, Kelly et al. 2010; Ahuja, Sharma et al. 2016).

The use of cleft-discordant sibling pairs for validation represents a methodological advance that addresses several limitations of traditional case-control studies. By comparing affected and unaffected siblings, we controlled for genetic background, shared environmental exposures, and population stratification while maintaining the power to detect disease-relevant epigenetic differences (Bell, Pai et al. 2011; Bell and Spector 2011; Bell and Saffery 2012; Bell and Spector 2012).

The integration of multiple analytical approaches, including meQTL mapping, differential methylation analysis, and functional annotation, provides convergent evidence for the biological significance of our findings (Villicaña and Bell 2021; Villicaña, Castillo-Fernandez et al. 2023). The high concordance (86%) between our disease-specific validation and population-level methylation effects in the mQTL Database demonstrates that OFC-associated genetic variants influence methylation through mechanisms that are conserved across different populations and study designs (Grundberg, Meduri et al. 2013; McRae, Powell et al. 2014).

Some limitations should be acknowledged in interpreting our findings. The cross-sectional nature of our methylation measurements does not capture the dynamic changes in epigenetic patterns that occur during development (Feinberg 2007; Relton and Davey Smith 2010). The tissue types analyzed (blood and saliva) may not fully represent the cell populations most relevant to craniofacial development, though they offer practical advantages for large-scale studies and clinical applications (Davies, Volta et al. 2012; Lowe, Gemma et al. 2013) and previous studies have shown high correlation with methylation levels in lip and palate (Alvizi, Ke et al. 2017; Sharp, Ho et al. 2017).

The functional consequences of the methylation changes that we identified remain to be demonstrated through direct experimental validation. While our bioinformatic analysis suggests these changes occur at regulatory elements (Fishilevich, Nudel et al. 2017; Wilderman, VanOudenhove et al. 2018; Yankee, Oh et al. 2023), functional studies using cellular or animal models will be necessary to confirm their effects on gene expression and developmental processes. The integration of our findings with single-cell epigenomics approaches could provide more detailed insights into the cell-type-specific effects of these genetic variants during craniofacial development.

Our findings have several potential clinical applications, especially for developing biomarkers for OFC risk assessment, particularly in families with existing genetic risk factors (Widschwendter, Fiegl et al. 2007; Teschendorff, Menon et al. 2010). The tissue-specific effects we observed suggest that saliva-based methylation testing might provide a non-invasive approach for risk stratification (Park, Park et al. 2011; Langevin, Koestler et al. 2012). However, the development of clinical applications would require validation in larger, more diverse populations.

In summary, our findings contribute to a more complete understanding of OFC etiology that could inform prevention strategies (Barker 2007; Gluckman, Hanson et al. 2008). The identification of environmental factors that influence the methylation patterns we characterized could lead to targeted interventions during pregnancy or early development. The transgenerational effects suggested by our 8q24 findings indicate that maternal health and environmental exposures might influence OFC risk in offspring through epigenetic mechanisms (Guo, Choufani et al. 2008; Joubert, den Dekker et al. 2016).

## Conclusions

This study provides a systematic characterization of meQTLs in orofacial clefts, offering mechanistic insights that bridge the gap between genetic associations and functional consequences. Our findings resolve the longstanding mystery surrounding the 8q24 locus by demonstrating its role in *MYC* regulation through long-range epigenetic mechanisms. The developmental timing of methylation effects, tissue-specific patterns of association, and convergent evidence from multiple analytical approaches support a model in which genetic variants influence OFC risk through modulation of DNA methylation at regulatory elements critical for craniofacial development.

These results demonstrate the power of integrating genetic and epigenetic analyses to understand complex disease etiology and highlight the importance of considering temporal and tissue-specific factors in studies of developmental disorders (Dunham, Kundaje et al. 2012; Roadmap Epigenomics, Kundaje et al. 2015). The methodological framework we established could be applied to other complex birth defects to identify functional mechanisms underlying genetic associations (Bernstein, Stamatoyannopoulos et al. 2010; Kundaje, Meuleman et al. 2015). Our findings provide a foundation for future functional studies and suggest potential avenues for therapeutic intervention targeting epigenetic mechanisms in OFC prevention and treatment (Bernstein, Stamatoyannopoulos et al. 2010; Stunnenberg and Hirst 2016).

## Data Availability

All data produced in the present study are available upon reasonable request to the authors.

